# Validation and Refinement of PreICH scale to Identify Intracerebral Hemorrhage Versus Large-Vessel Occlusion

**DOI:** 10.64898/2026.07.07.26357511

**Authors:** Albert Freixa, Gerard Mauri, Yhovany Gallego, Anna Garcia-Diaz, Cristina Nieva, Mikel Vicente-Pascual, Laura Perez-Girona, Eduardo San Pedro-Murillo, Eugenia Saureu-Rufach, Raquel Mitjana, Sara Salvany, Ares Peguera, Cristina Pereira, Francisco Purroy

**Affiliations:** Stroke Unit, Department of Neurology, Hospital Universitari Arnau de Vilanova de Lleida, Lleida, Spain; Clinical Neuroscience research group,Institut de Recerca Biomèdica de Lleida Fundació Dr. Pifarré (IRBLleida), Lleida, Spain; Departament de Medicina i Cirugia, Universitat de Lleida (UdL), Lleida, Spain; Department of Radiology, Hospital Universitari Arnau de Vilanova de Lleida, Lleida, Spain; Institut de Diagnostic per la Imatge, Lleida, Spain; Departament d’Infermeria i Fisioteràpia, Universitat de Lleida (UdL), Lleida, Spain

**Author notes:** Corresponding Author: Francisco Purroy, MD, PhD. Stroke Unit, Department of Neurology-Hospital Universitari Arnau de Vilanova de Lleida, Lleida, Spain.

**Keywords:** intracerebral hemorrhage, large-vessel occlusion, prehospital stroke triage, RACE, PreICH, clinical prediction model

## Abstract

**Background and Purpose:** -Prehospital large-vessel occlusion (LVO) scales identify severe stroke syndromes but may not distinguish LVO from intracerebral hemorrhage (ICH). We aimed to prospectively validate the PreICH scale, with the primary diagnostic objective of differentiating ICH from confirmed LVO, and to explore whether additional hemorrhage-oriented variables could refine its performance.

**Methods:** -We conducted a prospective observational study of consecutive stroke-code activations evaluated before neuroimaging by a vascular neurologist. PreICH was calculated prospectively. Patients with calculable PreICH and valid final diagnosis were included. The primary diagnostic cohort comprised confirmed LVO and ICH. Secondary cohorts included ischemic stroke versus ICH and the overall stroke-code cohort, including stroke mimics. Multivariable NIHSS-adjusted models identified variables associated with ICH. A modified PreICH score (mPreICH) was derived post hoc and evaluated as exploratory apparent performance.

**Results:** -Among 1012 screened activations, 982 patients were analyzed: 597 ischemic strokes, 91 ICH, and 294 stroke mimics. The LVO-versus-ICH cohort included 144 LVO and 91 ICH. NIHSS and RACE were higher in ICH than in ischemic stroke, but did not differ between LVO and ICH (NIHSS, 13 [IQR, 7-20] versus 15 [5-23], P=0.300; RACE, 5 [2-8] versus 6 [2-8], P=0.435). In the LVO-versus-ICH cohort, PreICH showed an AUC of 0.758 (95% CI, 0.696-0.820), whereas RACE did not discriminate LVO from ICH (AUC, 0.530 [95% CI, 0.453-0.607]). The exploratory mPreICH showed apparent AUCs of 0.835 (95% CI, 0.785-0.884) for ischemic stroke versus ICH and 0.798 (95% CI, 0.740-0.856) for LVO versus ICH.

**Conclusions:** -In this prospective stroke-code cohort, severity-based scales distinguished ICH from the overall ischemic stroke population but showed limited ability to differentiate LVO from ICH. An exploratory modified PreICH scale incorporating additional hemorrhage-oriented variables improved apparent discriminative performance, including in the LVO-versus-ICH setting. External validation is required before potential implementation in prehospital decision-making.

## Introduction

Prehospital stroke triage has become increasingly oriented toward the early identification of patients with suspected large-vessel occlusion (LVO), because direct transfer to thrombectomy-capable centers may reduce time to reperfusion and improve outcomes in appropriately selected patients.^1,2^ Several prehospital stroke severity scales have therefore been developed to identify patients with a higher probability of LVO by capturing severe motor and cortical stroke syndromes. In the PRESTO study, which prospectively compared 8 prehospital stroke scales, RACE, G-FAST, and CG-FAST showed the best performance for anterior-circulation LVO and approached the performance of clinician-assessed NIHSS.^3^ However, these scales were designed primarily to identify severe ischemic stroke syndromes rather than to distinguish LVO from intracerebral hemorrhage (ICH).

This distinction is clinically relevant because ICH can present with severe stroke syndromes that overlap substantially with LVO. Accordingly, ICH is a frequent cause of false-positive results in prehospital LVO screening.^5^ The consequences of this overlap are not merely diagnostic. Prehospital routing determines whether patients are transported to the closest stroke center or bypassed directly to a thrombectomy-capable center.^2^ A mothership strategy may benefit selected patients with true LVO, but it may delay diagnosis, blood pressure management, anticoagulation reversal, neurosurgical evaluation, and early hemorrhage-oriented care in patients ultimately found to have ICH.^6,7^ In a secondary analysis of the RACECAT randomized clinical trial, bypassing the closest stroke center was associated with reduced chances of functional independence at 90 days among patients with a final diagnosis of ICH.^8^ Although neuroimaging remains mandatory for definitive diagnosis, clinical assessment is often the first source of information available for prehospital triage and hospital pre-alert. Several clinical findings have been associated with a higher likelihood of ICH, including elevated blood pressure, headache, vomiting, seizures, and impaired consciousness.^11,12^ Recent systematic reviews of clinical signs and prehospital prediction models for ICH suggest that these variables may contribute diagnostic information, but also highlight important limitations of existing models, including retrospective designs, heterogeneous variable definitions, exclusion of stroke mimics, limited external validation, and risk of bias.^12,13^ Importantly, most previous models focused on distinguishing ICH from ischemic stroke overall and did not specifically evaluate the clinically challenging comparison between confirmed LVO and ICH.^13^

A hemorrhage-oriented clinical scale could complement, rather than replace, severity-based LVO screening tools. Such an approach should not delay acute stroke pathways or substitute for noncontrast CT, but it could refine pre-alert communication, improve emergency department preparedness, and help characterize patients in whom a hemorrhagic phenotype is more likely. This may be particularly relevant in regional stroke systems where transport times, access to thrombectomy-capable centers, and the potential consequences of bypassing the closest stroke center vary substantially.^2,8^ Early recognition of a hemorrhage-enriched phenotype may also be relevant for time-sensitive aspects of acute care, including blood pressure management, anticoagulation reversal, neurocritical care, and neurosurgical preparedness.^6,7^

We previously proposed the PreICH scale as a clinical score designed to support early identification of ICH using readily available variables in a stroke-code setting, including systolic blood pressure >160 mm Hg, headache, male sex, coma, and hypercholesterolemia.^15^ In the present study, we aimed to prospectively validate the original PreICH scale in a consecutive stroke-code cohort, with the primary diagnostic objective of distinguishing ICH from confirmed LVO, the key thrombectomy-era differential diagnosis for acute stroke triage. Secondary objectives were to evaluate PreICH performance in the broader ischemic stroke-versus-ICH comparison, in the overall stroke-code population including stroke mimics, and across predefined RACE strata. In addition, based on previous evidence indicating that vomiting, seizure, impaired consciousness, headache, and elevated blood pressure increase the likelihood of hemorrhagic stroke,^11,12^ we prospectively assessed hemorrhage-oriented variables not included in the original scale to explore whether these variables could support an exploratory refinement of PreICH.

## Methods

### Study Design, Setting, and Ethics

We conducted a prospective observational study including consecutive stroke-code activations evaluated at the Emergency Department of Hospital Universitari Arnau de Vilanova, Lleida, Spain, between May 2024 and October 2025. Patients were managed according to the institutional acute stroke protocol. Stroke-code activations were performed according to the Stroke Plan of the Government of Catalonia. Stroke code could be activated in the prehospital setting, with subsequent transfer to our hospital, or at hospital arrival by the emergency medical services or emergency department team. In general, patients with clinical suspicion of acute stroke within 24 hours from symptom onset or last time known well, and with prestroke functional status compatible with acute stroke-code activation, were eligible for evaluation.

The study was approved by the local Research Ethics Committee of Hospital Universitari Arnau de Vilanova de Lleida (Comite d’Etica i Investigacio Clinica; ID: CEIC 3220). All participants or their legal representatives received study information, and written informed consent was obtained before participation. The study was conducted in accordance with relevant guidelines and regulations, including Good Clinical Practice principles. The processing, communication, and transfer of personal data complied with Spanish Organic Law 3/2018 on Personal Data Protection and Guarantee of Digital Rights and Regulation (EU) 2016/679 of the European Parliament and of the Council. All statistical analyses were performed on an anonymized dataset. The manuscript was prepared following the Strengthening the Reporting of Observational Studies in Epidemiology (STROBE) reporting recommendations for observational studies.

### Study Population

Patients were eligible for inclusion if they met regional stroke-code activation criteria. Patients were included irrespective of final diagnosis. Patients without the clinical variables required to calculate PreICH or without valid final diagnostic classification were excluded from the analytic cohort.

Three analytic cohorts were defined. The overall stroke-code cohort comprised all patients with calculable PreICH and valid final diagnostic classification, including ischemic stroke, ICH, and stroke mimics. The ischemic stroke-versus-ICH cohort comprised all patients with a final diagnosis of ischemic stroke or ICH and was used to contextualize PreICH performance across confirmed stroke diagnoses. The LVO-versus-ICH cohort comprised patients with confirmed LVO or ICH and was used for the primary diagnostic comparison, addressing the clinically relevant thrombectomy-era distinction between severe ischemic stroke requiring consideration of endovascular pathways and hemorrhagic stroke requiring hemorrhage-oriented care.

### Clinical Assessment and Data Collection

All patients were evaluated at hospital arrival by a vascular neurologist responsible for the emergency stroke-code assessment. Clinical variables were prospectively recorded by the vascular neurologist during the initial stroke-code evaluation and before any neuroimaging was performed. Recorded variables included demographic characteristics, vascular risk factors, history of atrial fibrillation, prior stroke or transient ischemic attack, baseline modified Rankin Scale score, previous medication use, systolic and diastolic blood pressure, headache at onset, nausea or vomiting, seizure at onset, Glasgow Coma Scale, level of consciousness, NIHSS, RACE, and PreICH scores.

Vascular risk factors, previous treatments, and medical history were obtained from available electronic medical records, patient or witness interview, and emergency medical documentation. Definitions of vascular risk factors were consistent with those used in the original PreICH derivation study.^15^ Hypertension was defined as prior diagnosis, systolic blood pressure >=140 mm Hg, diastolic blood pressure >=90 mm Hg, or current use of antihypertensive medication. Diabetes mellitus was defined as a history of fasting glucose >=126 mg/dL or current use of glucose-lowering medication. Hypercholesterolemia was defined as total cholesterol >=220 mg/dL or current use of lipid-lowering therapy. Current or recent smoking was defined as cigarette smoking during the previous 5 years. Alcoholism was defined as a prior clinical diagnosis of alcohol use disorder, alcohol dependence, or harmful alcohol consumption documented in the medical record or reported during the initial clinical assessment.

History of coronary artery disease, peripheral arterial disease, atrial fibrillation, valvular heart disease, prior stroke or transient ischemic attack, chronic kidney disease, and cognitive impairment was also recorded.

PreICH was calculated prospectively at the time of the initial patient evaluation according to the original score definition. The RACE score was calculated at hospital arrival before neuroimaging, with individualized prospective recording of each RACE item, according to the original RACE scale description.^17^ Glasgow Coma Scale was also calculated during the same initial preimaging clinical assessment. Blood pressure was defined according to the first available measurement obtained during the earliest patient assessment. Headache, nausea/vomiting, and seizure at onset were recorded when present at symptom onset or during the initial stroke-code assessment. Seizure at onset was defined as a witnessed or clinically reported epileptic seizure accompanying or immediately preceding the neurological deficit. Level of consciousness was categorized into 4 levels according to the NIHSS level-of-consciousness item: alert, somnolent with response to verbal commands, obtunded with response to painful stimulus, and coma.

For stratified analyses, RACE was dichotomized using the conventional threshold of >4 points, with patients classified as RACE <=4 or RACE >4.^17^ After prospective data collection, the database was systematically reviewed to verify completeness, internal consistency, and final diagnostic classification.

### Neuroimaging Protocol and Diagnostic Classification

All patients underwent multimodal CT according to the institutional stroke-code imaging protocol. The acute imaging protocol included noncontrast brain CT, CT angiography, and CT perfusion. Imaging data were initially processed using RAPID/RapidAI software (iSchemaView, Inc., Menlo Park, CA, USA) and subsequently reviewed and validated by 2 experienced neuroradiologists.

Neuroimaging was used to identify ICH, confirm ischemic stroke, detect LVO, and support final diagnostic classification. LVO was defined according to the original PreICH study and prior work using a broad definition of occlusion site, as an occlusion of the intracranial internal carotid artery, carotid terminus, M1 or proximal M2 segment of the middle cerebral artery, basilar artery, or P1 segment of the posterior cerebral artery on CT angiography or subsequent vascular imaging review.^15,16^ Final diagnosis was established after completion of the acute hospital evaluation, integrating neurological assessment, neuroimaging findings, and subsequent diagnostic information when required.

### PreICH Validation and Exploratory Refinement

The original PreICH scale was calculated prospectively during the initial stroke-code assessment and is referred to throughout the manuscript as PreICH. PreICH was evaluated as an ordinal score and by receiver operating characteristic curve analysis to assess its discriminative performance for identifying ICH, with the LVO-versus-ICH cohort considered the primary diagnostic comparison.

In addition to variables included in the original PreICH framework, nausea/vomiting, seizure at onset, and graded impairment of consciousness were prospectively recorded because of their biological plausibility and their reported association with hemorrhagic stroke. These variables were not used to define the original PreICH validation analysis.

After completion of the primary PreICH validation analyses, multivariable logistic regression adjusted for NIHSS was performed to identify variables independently associated with ICH. These results were then used to explore whether the original PreICH scale could be refined to improve hemorrhage-oriented discrimination. Any modified version of PreICH derived from these analyses was considered exploratory, calculated post hoc, and not a prespecified validation target.

### Outcomes

The primary diagnostic outcome was final diagnosis of ICH versus confirmed LVO in the LVO-versus-ICH cohort. Secondary diagnostic outcomes were ICH versus ischemic stroke in the broader ischemic stroke-versus-ICH cohort and ICH versus nonhemorrhagic diagnosis in the overall stroke-code cohort. Additional analyses evaluated diagnostic performance according to predefined RACE strata. ICH was considered the event of interest in all diagnostic performance analyses.

### Sample Size

The target sample size was informed by the original PreICH derivation study, in which 989 stroke-code activations were screened, 285 patients with either LVO or ICH were included, and PreICH could be calculated in 281 patients. In that study, the prespecified sample size calculation estimated that at least 252 patients were required for the LVO-versus-ICH comparison. Therefore, for the present prospective validation study, we aimed to include at least 1000 consecutive stroke-code activations during the predefined study period to obtain a cohort comparable to the original derivation study and to support the primary diagnostic and secondary analyses.

### Statistical Analysis

Continuous variables were evaluated for normality using visual inspection and summary statistics. Normally distributed continuous variables are reported as mean+-SD and compared using the Student t test for 2-group comparisons or 1-way analysis of variance for comparisons across more than 2 groups. Non-normally distributed continuous variables and ordinal variables, including NIHSS, RACE, PreICH, modified PreICH (mPreICH), Glasgow Coma Scale, and baseline modified Rankin Scale score, are reported as median with interquartile range and compared using the Mann-Whitney U test or Kruskal-Wallis test, as appropriate. Categorical variables are reported as counts and percentages and compared using the chi-square test or Fisher exact test when expected cell counts were <5.

No imputation was performed. After exclusion of patients with insufficient data to calculate PreICH or invalid final diagnostic classification, analyses were performed on complete available data for each analytic cohort.

Multivariable logistic regression was performed to identify variables independently associated with ICH. NIHSS was included as an adjustment variable because overall neurological severity may confound the association between individual clinical features and ICH diagnosis. Two separate models were fitted: one in the primary LVO-versus-ICH cohort and one in the broader ischemic stroke-versus-ICH cohort. Candidate variables were selected based on the original PreICH components and prior evidence supporting their association with hemorrhagic stroke, rather than on univariable screening alone. These variables included NIHSS, systolic blood pressure >160 mm Hg, headache, sex, hypercholesterolemia and/or lipid-lowering treatment, nausea/vomiting, seizure at onset, and level of consciousness. After review of descriptive comparisons, diabetes mellitus and anticoagulation were not considered for model-based refinement because they did not differentiate the primary LVO-versus-ICH cohort and were not part of the original PreICH framework or the predefined hemorrhage-oriented clinical construct. Level of consciousness was entered as a categorical variable, with alert status as the reference category. Results are reported as adjusted odds ratios with 95% confidence intervals and P values.

The discriminative performance of PreICH and RACE was assessed using receiver operating characteristic curve analysis. Areas under the curve with 95% confidence intervals were estimated in the overall stroke-code cohort, the ischemic stroke-versus-ICH cohort, the primary LVO-versus-ICH cohort, and across predefined RACE strata. After review of the NIHSS-adjusted multivariable models, an exploratory post hoc modified PreICH score (mPreICH) was calculated to assess whether hemorrhage-oriented variables associated with ICH could improve apparent discrimination. The discriminative performance of mPreICH was evaluated using receiver operating characteristic curve analysis in the same analytic cohorts. Because mPreICH was derived and evaluated within the same dataset, these analyses were considered exploratory and reflected apparent performance within the derivation dataset.

Threshold-specific performance of mPreICH was evaluated using candidate cutoffs. Sensitivity, specificity, positive predictive value, negative predictive value, positive likelihood ratio, negative likelihood ratio, and Youden index were calculated in the ischemic stroke-versus-ICH cohort and in the primary LVO-versus-ICH cohort. These analyses were exploratory and were not intended to define definitive triage thresholds. All statistical tests were 2-sided, and P<0.05 was considered statistically significant. Statistical analyses were performed using IBM SPSS Statistics, version 29.0.

## Results

### Study Population

Between May 2024 and October 2025, 1012 consecutive stroke-code activations were screened. Thirty patients were excluded because of insufficient data to calculate PreICH or invalid final diagnostic classification. The final overall stroke-code cohort included 982 patients: 597 with ischemic stroke (60.8%), 91 with intracerebral hemorrhage (ICH; 9.3%), and 294 with stroke mimics (29.9%). Baseline and early clinical characteristics of the overall cohort are shown in Supplemental Table 1. In the overall stroke-code cohort, patients with ICH had greater neurological severity than those with ischemic stroke or stroke mimics, as reflected by higher NIHSS and RACE scores and lower Glasgow Coma Scale scores. ICH was also associated with higher systolic blood pressure and more frequent hemorrhage-oriented clinical features, including headache, nausea or vomiting, and severe impairment of consciousness. Stroke mimics were younger and had lower median NIHSS and RACE scores than patients with ischemic stroke or ICH.

The ischemic stroke-versus-ICH cohort included 688 patients: 597 with ischemic stroke (86.8%) and 91 with ICH (13.2%). Among patients with ischemic stroke, 144 had confirmed large-vessel occlusion (LVO), representing 24.1% of ischemic strokes and 20.9% of the ischemic stroke-versus-ICH cohort. The LVO-versus-ICH cohort, used for the primary diagnostic comparison, included 235 patients: 144 with confirmed LVO (61.3%) and 91 with ICH (38.7%).

### Ischemic Stroke Versus Intracerebral Hemorrhage Cohort

Clinical characteristics of patients with ischemic stroke and ICH are shown in Table 1. Age and sex distribution were similar between groups. Patients with ICH had higher systolic blood pressure than patients with ischemic stroke (168.9±33.0 versus 151.3±27.1 mm Hg; P<0.001), and systolic blood pressure >160 mm Hg was more frequent in ICH (53/91 [58.2%] versus 205/597 [34.3%]; P<0.001). Diastolic blood pressure was also higher in ICH (97.7±22.6 versus 84.9±18.7 mm Hg; P<0.001). Stroke severity scales were higher in ICH than in ischemic stroke. Median NIHSS was 15 (IQR, 5-23) in ICH and 4 (IQR, 1-8) in ischemic stroke (P<0.001). Median RACE was 6 (IQR, 2-8) in ICH and 2 (IQR, 0-3) in ischemic stroke (P<0.001). Median PreICH was also higher in ICH than in ischemic stroke (2 [IQR, 1-3] versus 1 [IQR, 0-2]; P<0.001; Table 1).

**Table 1.** Clinical Characteristics of the Ischemic Stroke-Versus-ICH Cohort.

| Characteristic | Total, N=688 | Ischemic Stroke, N=597 | ICH, N=91 | P Value |
| --- | --- | --- | --- | --- |
| **Demographics** |  |  |  |  |
| Age, y, mean±SD | 72.0±14.3 | 72.2±14.0 | 70.9±16.6 | 0.413 |
| Female sex | 304 (44.2) | 261 (43.7) | 43 (47.3) | 0.527 |
| Male sex | 384 (55.8) | 336 (56.3) | 48 (52.7) | 0.527 |
| **Blood pressure** |  |  |  |  |
| Systolic blood pressure, mm Hg, mean±SD | 153.6±28.6 | 151.3±27.1 | 168.9±33.0 | <0.001 |
| Diastolic blood pressure, mm Hg, mean±SD | 86.6±19.7 | 84.9±18.7 | 97.7±22.6 | <0.001 |
| Systolic blood pressure >160 mm Hg | 258 (37.5) | 205 (34.3) | 53 (58.2) | <0.001 |
| **Clinical scales** |  |  |  |  |
| NIHSS, median (IQR) | 4 (2–11) | 4 (1–8) | 15 (5–23) | <0.001 |
| RACE, median (IQR) | 2 (0–4) | 2 (0–3) | 6 (2–8) | <0.001 |
| PreICH, median (IQR) | 1 (0–2) | 1 (0–2) | 2 (1–3) | <0.001 |
| mPreICH, median (IQR) | 2 (0–2) | 0 (0–2) | 4 (2–5) | <0.001 |
| Glasgow Coma Scale, median (IQR) | 15 (15–15) | 15 (15–15) | 14 (10–15) | <0.001 |
| Baseline mRS, median (IQR) | 1 (0–1) | 1 (0–1) | 1 (0–2) | 1.000 |
| **Neuroimaging** |  |  |  |  |
| Confirmed LVO | 144 (20.9) | 144 (24.1) | 0 (0.0) | — |
| No confirmed LVO | 544 (79.1) | 453 (75.9) | 91 (100.0) | — |
| **Vascular risk factors and medical history** |  |  |  |  |
| Hypertension | 484 (70.3) | 424 (71.0) | 60 (65.9) | 0.322 |
| Dyslipidemia | 368 (53.5) | 325 (54.4) | 43 (47.3) | 0.200 |
| Diabetes mellitus | 212 (30.8) | 193 (32.3) | 19 (20.9) | 0.028 |
| Previous atrial fibrillation | 104 (15.1) | 82 (13.7) | 22 (24.2) | 0.033 |
| Previous TIA or ischemic stroke | 98 (14.2) | 94 (15.7) | 4 (4.4) | 0.004 |
| Alcoholism | 60 (8.7) | 56 (9.4) | 4 (4.4) | 0.268 |
| Smoking | 99 (14.4) | 91 (15.2) | 8 (8.8) | 0.259 |
| **Previous treatments** |  |  |  |  |
| Statins | 293 (42.6) | 262 (43.9) | 31 (34.1) | 0.191 |
| Antiplatelet treatment | 161 (23.4) | 151 (25.3) | 10 (11.0) | 0.010 |
| Anticoagulation | 108 (15.7) | 85 (14.2) | 23 (25.3) | 0.007 |
| Oral antidiabetic drugs | 183 (26.6) | 170 (28.5) | 13 (14.3) | 0.015 |
| ACE inhibitors | 231 (33.6) | 209 (35.0) | 22 (24.2) | 0.042 |
| Angiotensin receptor blockers | 122 (17.7) | 107 (17.9) | 15 (16.5) | 0.738 |
| Diuretics | 210 (30.5) | 187 (31.3) | 23 (25.3) | 0.243 |
| **Symptoms and early clinical presentation** |  |  |  |  |
| Headache | 88 (12.8) | 59 (9.9) | 29 (31.9) | <0.001 |
| Nausea or vomiting | 62 (9.0) | 38 (6.4) | 24 (26.4) | <0.001 |
| Severe impairment of consciousness, GCS <8 | 20 (2.9) | 4 (0.7) | 16 (17.6) | <0.001 |
| Level of consciousness |  |  |  | <0.001 |
| Alert | 585 (85.0) | 541 (90.6) | 44 (48.4) |  |
| Somnolent, responds to verbal commands | 62 (9.0) | 39 (6.5) | 23 (25.3) |  |
| Obtunded, responds to painful stimulus | 19 (2.8) | 10 (1.7) | 9 (9.9) |  |
| Coma | 22 (3.2) | 7 (1.2) | 15 (16.5) |  |

Values are n (%) unless otherwise indicated. P values compare ischemic stroke versus ICH. P values are not shown for confirmed LVO because this variable is part of the diagnostic characterization of ischemic stroke and defines the LVO-versus-ICH cohort. ACE indicates angiotensin-converting enzyme; GCS, Glasgow Coma Scale; ICH, intracerebral hemorrhage; IQR, interquartile range; LVO, large-vessel occlusion; mPreICH, modified PreICH; mRS, modified Rankin Scale; NIHSS, National Institutes of Health Stroke Scale; RACE, Rapid Arterial occlusion Evaluation; SD, standard deviation; and TIA, transient ischemic attack.

Several hemorrhage-oriented clinical features were more frequent in ICH than in ischemic stroke. Headache was present in 29 patients with ICH (31.9%) and 59 patients with ischemic stroke (9.9%; P<0.001). Nausea or vomiting was present in 24 patients with ICH (26.4%) and 38 patients with ischemic stroke (6.4%; P<0.001). Severe impairment of consciousness, defined as Glasgow Coma Scale <8, was observed in 16 patients with ICH (17.6%) and 4 patients with ischemic stroke (0.7%; P<0.001). Level of consciousness differed significantly between groups: 44 patients with ICH (48.4%) were alert compared with 541 patients with ischemic stroke (90.6%), whereas coma was present in 15 patients with ICH (16.5%) and 7 patients with ischemic stroke (1.2%; P<0.001; Table 1).

Regarding vascular risk factors and previous treatments, diabetes mellitus, previous transient ischemic attack or ischemic stroke, antiplatelet treatment, oral antidiabetic drugs, and angiotensin-converting enzyme inhibitor use were more frequent in ischemic stroke, whereas previous atrial fibrillation and anticoagulation were more frequent in ICH (Table 1).

Individual RACE components are shown in Supplemental Table 2. In the overall stroke-code cohort and in the ischemic stroke-versus-ICH cohort, all RACE components differed significantly across diagnostic groups. In the ischemic stroke-versus-ICH cohort, ICH was associated with more frequent moderate/severe facial palsy, severe arm paresis, severe leg paresis, gaze deviation, agnosia, and inability to obey commands (Supplemental Table 2).

### LVO-Versus-ICH Cohort

Clinical characteristics of patients with confirmed LVO and ICH are shown in Table 2. In contrast to the comparison with the overall ischemic stroke group, NIHSS and RACE did not significantly differ between LVO and ICH. Median NIHSS was 13 (IQR, 7-20) in LVO and 15 (IQR, 5-23) in ICH (P=0.300). Median RACE was 5 (IQR, 2-8) in LVO and 6 (IQR, 2-8) in ICH (P=0.435; Table 2). In the RACE-component analysis, facial palsy, gaze deviation, agnosia, and the commands/language item did not differ between LVO and ICH, whereas arm and leg paresis differed modestly (Supplemental Table 2). Despite similar NIHSS and RACE scores, several hemorrhage-oriented variables continued to distinguish ICH from LVO. Patients with ICH had higher systolic blood pressure than patients with LVO (168.9±33.0 versus 145.9±22.8 mm Hg; P<0.001), and systolic blood pressure >160 mm Hg was more frequent in ICH (53/91 [58.2%] versus 35/144 [24.3%]; P<0.001). Headache was more frequent in ICH than in LVO (29/91 [31.9%] versus 11/144 [7.6%]; P<0.001), as was nausea or vomiting (24/91 [26.4%] versus 6/144 [4.2%]; P<0.001). Severe impairment of consciousness was also more frequent in ICH (16/91 [17.6%] versus 3/144 [2.1%]; P<0.001), and level of consciousness differed significantly between groups (P<0.001; Table 2).

**Table 2.** Clinical Characteristics of the LVO-Versus-ICH Cohort.

| Characteristic | Total, N=235 | LVO, N=144 | ICH, N=91 | P Value |
| --- | --- | --- | --- | --- |
| **Demographics** |  |  |  |  |
| Age, y, mean±SD | 72.7±15.2 | 73.9±14.2 | 70.9±16.6 | 0.138 |
| Female sex | 120 (51.1) | 77 (53.5) | 43 (47.3) | 0.353 |
| Male sex | 115 (48.9) | 67 (46.5) | 48 (52.7) | 0.353 |
| **Blood pressure** |  |  |  |  |
| Systolic blood pressure, mm Hg, mean±SD | 154.8±29.4 | 145.9±22.8 | 168.9±33.0 | <0.001 |
| Diastolic blood pressure, mm Hg, mean±SD | 88.2±20.6 | 82.1±16.7 | 97.7±22.6 | <0.001 |
| Systolic blood pressure >160 mm Hg | 88 (37.4) | 35 (24.3) | 53 (58.2) | <0.001 |
| **Clinical scales** |  |  |  |  |
| NIHSS, median (IQR) | 13 (7–21) | 13 (7–20) | 15 (5–23) | 0.300 |
| RACE, median (IQR) | 5 (2–8) | 5 (2–8) | 6 (2–8) | 0.435 |
| PreICH, median (IQR) | 1 (0–2) | 1 (0–1.8) | 2 (1–3) | <0.001 |
| mPreICH, median (IQR) | 2 (0–4) | 0 (0–2) | 4 (2–5) | <0.001 |
| Glasgow Coma Scale, median (IQR) | 15 (14–15) | 15 (14–15) | 14 (10–15) | <0.001 |
| Baseline mRS, median (IQR) | 0 (0–2) | 0 (0–1) | 1 (0–2) | 0.520 |
| **Vascular risk factors and medical history** |  |  |  |  |
| Hypertension | 161 (68.5) | 101 (70.1) | 60 (65.9) | 0.499 |
| Dyslipidemia | 117 (49.8) | 74 (51.4) | 43 (47.3) | 0.537 |
| Diabetes mellitus | 58 (24.7) | 39 (27.1) | 19 (20.9) | 0.283 |
| Previous atrial fibrillation | 53 (22.6) | 31 (21.5) | 22 (24.2) | 0.659 |
| Previous TIA or ischemic stroke | 24 (10.2) | 20 (13.9) | 4 (4.4) | 0.019 |
| Alcoholism | 10 (4.3) | 6 (4.2) | 4 (4.4) | 0.933 |
| Smoking | 23 (9.8) | 15 (10.4) | 8 (8.8) | 0.420 |
| **Previous treatments** |  |  |  |  |
| Statins | 85 (36.2) | 54 (37.5) | 31 (34.1) | 0.594 |
| Antiplatelet treatment | 42 (17.9) | 32 (22.2) | 10 (11.0) | 0.029 |
| Anticoagulation | 50 (21.3) | 27 (18.8) | 23 (25.3) | 0.234 |
| Oral antidiabetic drugs | 46 (19.6) | 33 (22.9) | 13 (14.3) | 0.104 |
| ACE inhibitors | 68 (28.9) | 46 (31.9) | 22 (24.2) | 0.201 |
| Angiotensin receptor blockers | 40 (17.0) | 25 (17.4) | 15 (16.5) | 0.862 |
| Diuretics | 81 (34.5) | 58 (40.3) | 23 (25.3) | 0.018 |
| **Symptoms and early clinical presentation** |  |  |  |  |
| Headache | 40 (17.0) | 11 (7.6) | 29 (31.9) | <0.001 |
| Nausea or vomiting | 30 (12.8) | 6 (4.2) | 24 (26.4) | <0.001 |
| Severe impairment of consciousness, GCS <8 | 19 (8.1) | 3 (2.1) | 16 (17.6) | <0.001 |
| Level of consciousness |  |  |  | <0.001 |
| Alert | 148 (63.0) | 104 (72.2) | 44 (48.4) |  |
| Somnolent, responds to verbal commands | 48 (20.4) | 25 (17.4) | 23 (25.3) |  |
| Obtunded, responds to painful stimulus | 18 (7.7) | 9 (6.3) | 9 (9.9) |  |
| Coma | 21 (8.9) | 6 (4.2) | 15 (16.5) |  |

Values are n (%) unless otherwise indicated. P values compare LVO versus ICH. LVO defines the ischemic stroke subgroup included in this primary diagnostic cohort. ACE indicates angiotensin-converting enzyme; GCS, Glasgow Coma Scale; ICH, intracerebral hemorrhage; IQR, interquartile range; LVO, large-vessel occlusion; mPreICH, modified PreICH; mRS, modified Rankin Scale; NIHSS, National Institutes of Health Stroke Scale; RACE, Rapid Arterial occlusion Evaluation; SD, standard deviation; and TIA, transient ischemic attack.

PreICH was higher in ICH than in LVO. Median PreICH was 2 (IQR, 1-3) in ICH and 1 (IQR, 0-1.8) in LVO (P<0.001; Table 2). Diabetes mellitus did not differ significantly between LVO and ICH (39/144 [27.1%] versus 19/91 [20.9%]; P=0.283), nor did anticoagulation (27/144 [18.8%] versus 23/91 [25.3%]; P=0.234; Table 2).

### Multivariable Models

In the primary LVO-versus-ICH model, NIHSS was not associated with ICH (adjusted odds ratio [OR] per 1-point increase, 0.985 [95% CI, 0.935-1.037]; P=0.564). Systolic blood pressure >160 mm Hg (adjusted OR, 2.334 [95% CI, 1.661-3.279]; P<0.001), headache (adjusted OR, 5.938 [95% CI, 2.386-14.773]; P<0.001), and nausea or vomiting (adjusted OR, 2.244 [95% CI, 1.295-3.890]; P=0.004) were independently associated with ICH. Level of consciousness was associated with ICH in the overall categorical test (P=0.017), with coma showing the strongest association (adjusted OR, 14.938 [95% CI, 2.792-79.930]; P=0.002; Table 3).

**Table 3.** NIHSS-Adjusted Multivariable Predictors of Intracerebral Hemorrhage.

| Predictor | LVO vs ICH Adjusted OR (95% CI) | P Value | Ischemic Stroke vs ICH Adjusted OR (95% CI) | P Value |
| --- | --- | --- | --- | --- |
| NIHSS, per 1-point increase | 0.985 (0.935–1.037) | 0.564 | 1.103 (1.056–1.152) | <0.001 |
| Systolic blood pressure >160 mm Hg | 2.334 (1.661–3.279) | <0.001 | 1.973 (1.477–2.634) | <0.001 |
| Headache | 5.938 (2.386–14.773) | <0.001 | 6.028 (3.087–11.770) | <0.001 |
| Male sex | 1.260 (0.657–2.419) | 0.486 | 0.969 (0.558–1.684) | 0.912 |
| Hypercholesterolemia/lipid-lowering treatment | 1.004 (0.484–2.085) | 0.991 | 0.712 (0.395–1.281) | 0.256 |
| Nausea or vomiting | 2.244 (1.295–3.890) | 0.004 | 1.760 (1.217–2.545) | 0.003 |
| Seizure at onset | 2.270 (0.646–7.978) | 0.201 | 5.242 (1.257–21.867) | 0.023 |
| Level of consciousness, overall | — | 0.017 | — | 0.128 |
| Somnolence vs alert | 1.89 (0.713–4.338) | 0.220 | 2.442 (1.087–5.490) | 0.031 |
| Obtundation vs alert | 3.558 (0.893–14.183) | 0.072 | 2.960 (0.789–11.113) | 0.108 |
| Coma vs alert | 14.938 (2.792–79.930) | 0.002 | 3.591 (0.770–16.741) | 0.104 |

Values are adjusted odds ratios with 95% CIs. ICH was the event of interest in all models. Level of consciousness was entered as a categorical variable, with alert status as the reference category. The overall P value corresponds to the global test for the level-of-consciousness variable. CI indicates confidence interval; ICH, intracerebral hemorrhage; LVO, large-vessel occlusion; NIHSS, National Institutes of Health Stroke Scale; and OR, odds ratio.

In the broader ischemic stroke-versus-ICH model, NIHSS was independently associated with ICH (adjusted OR per 1-point increase, 1.103 [95% CI, 1.056-1.152]; P<0.001). Systolic blood pressure >160 mm Hg (adjusted OR, 1.973 [95% CI, 1.477-2.634]; P<0.001), headache (adjusted OR, 6.028 [95% CI, 3.087-11.770]; P<0.001), nausea or vomiting (adjusted OR, 1.760 [95% CI, 1.217-2.545]; P=0.003), and seizure at onset (adjusted OR, 5.242 [95% CI, 1.257-21.867]; P=0.023) were independently associated with ICH. Somnolence was also associated with ICH compared with alert status (adjusted OR, 2.442 [95% CI, 1.087-5.490]; P=0.031). Male sex and hypercholesterolemia/lipid-lowering treatment were not independently associated with ICH in either model (Table 3).

### Exploratory Refinement of PreICH

After completion of the primary validation analyses and review of the NIHSS-adjusted multivariable models, an exploratory refined version of the score was generated and named modified PreICH (mPreICH). The rationale for this refinement was to preserve the hemorrhage-oriented clinical structure of PreICH while incorporating additional variables associated with ICH and replacing the original binary severe-consciousness item with a graded assessment of level of consciousness.

The final mPreICH assigned +2 points for systolic blood pressure >160 mm Hg, +1 point for headache, +2 points for somnolence, +3 points for obtundation, +4 points for coma, +2 points for nausea or vomiting, and +2 points for seizure at onset. Male sex and hypercholesterolemia/lipid-lowering treatment were not included in mPreICH because these variables were not independently associated with ICH in the NIHSS-adjusted models. Diabetes mellitus and anticoagulation were also not incorporated into the exploratory score because, although they differed in the ischemic stroke-versus-ICH descriptive comparison, they did not differ significantly in the clinically relevant LVO-versus-ICH comparison and were not part of the original PreICH framework or the predefined hemorrhage-oriented clinical construct. The components and point allocation of PreICH and mPreICH are shown in Supplemental Table 4. Because mPreICH was derived after review of the multivariable results and evaluated in the same dataset, its performance should be interpreted as exploratory and as reflecting apparent performance within the derivation dataset.

After derivation, mPreICH scores were higher in ICH than in ischemic stroke in the ischemic stroke-versus-ICH cohort (4 [IQR, 2-5] versus 0 [IQR, 0-2]; P<0.001; Table 1). In the LVO-versus-ICH cohort, mPreICH was also higher in ICH than in LVO (4 [IQR, 2-5] versus 0 [IQR, 0-2]; P<0.001; Table 2).

### Discriminative Performance of RACE, PreICH, and mPreICH

The discriminative performance of RACE, PreICH, and mPreICH is shown in Table 4. In the primary LVO-versus-ICH cohort, RACE did not discriminate LVO from ICH (AUC, 0.530 [95% CI, 0.453-0.607]; P=0.446). In contrast, PreICH showed an AUC of 0.758 (95% CI, 0.696-0.820; P<0.001), and mPreICH showed an apparent AUC of 0.798 (95% CI, 0.740-0.856; P<0.001; Table 4).

**Table 4.** Discriminative Ability of RACE, PreICH, and mPreICH for Identifying Intracerebral Hemorrhage.

| Analysis Cohort | RACE Stratum | Score | AUC | SE | 95% CI | P Value |
| --- | --- | --- | --- | --- | --- | --- |
| Overall ROC analysis |  |  |  |  |  |  |
| Overall cohort | All patients | RACE | 0.774 | 0.028 | 0.719–0.828 | <0.001 |
| Overall cohort | All patients | PreICH | 0.717 | 0.026 | 0.665–0.768 | <0.001 |
| Overall cohort | All patients | mPreICH | 0.812 | 0.025 | 0.763–0.861 | <0.001 |
| Ischemic stroke vs ICH | All patients | RACE | 0.746 | 0.030 | 0.687–0.805 | <0.001 |
| Ischemic stroke vs ICH | All patients | PreICH | 0.714 | 0.027 | 0.661–0.768 | <0.001 |
| Ischemic stroke vs ICH | All patients | mPreICH | 0.835 | 0.025 | 0.785–0.884 | <0.001 |
| LVO vs ICH | All patients | RACE | 0.530 | 0.039 | 0.453–0.607 | 0.446 |
| LVO vs ICH | All patients | PreICH | 0.758 | 0.032 | 0.696–0.820 | <0.001 |
| LVO vs ICH | All patients | mPreICH | 0.798 | 0.030 | 0.740–0.856 | <0.001 |
| RACE-stratified ROC analysis |  |  |  |  |  |  |
| Overall cohort | RACE ≤4 | PreICH | 0.738 | 0.033 | 0.672–0.804 | <0.001 |
| Overall cohort | RACE ≤4 | mPreICH | 0.726 | 0.044 | 0.640–0.811 | <0.001 |
| Overall cohort | RACE >4 | PreICH | 0.734 | 0.040 | 0.655–0.812 | <0.001 |
| Overall cohort | RACE >4 | mPreICH | 0.768 | 0.037 | 0.696–0.841 | <0.001 |
| Ischemic stroke vs ICH | RACE ≤4 | PreICH | 0.730 | 0.035 | 0.662–0.799 | <0.001 |
| Ischemic stroke vs ICH | RACE ≤4 | mPreICH | 0.749 | 0.044 | 0.662–0.836 | <0.001 |
| Ischemic stroke vs ICH | RACE >4 | PreICH | 0.748 | 0.041 | 0.668–0.828 | <0.001 |
| Ischemic stroke vs ICH | RACE >4 | mPreICH | 0.829 | 0.034 | 0.762–0.897 | <0.001 |
| LVO vs ICH | RACE ≤4 | PreICH | 0.773 | 0.045 | 0.685–0.861 | <0.001 |
| LVO vs ICH | RACE ≤4 | mPreICH | 0.779 | 0.048 | 0.684–0.874 | <0.001 |
| LVO vs ICH | RACE >4 | PreICH | 0.752 | 0.043 | 0.667–0.836 | <0.001 |
| LVO vs ICH | RACE >4 | mPreICH | 0.821 | 0.037 | 0.749–0.893 | <0.001 |

AUC indicates area under the receiver operating characteristic curve; CI, confidence interval; ICH, intracerebral hemorrhage; LVO, large-vessel occlusion; mPreICH, modified PreICH; RACE, Rapid Arterial occlusion Evaluation; and SE, standard error.

In secondary analyses, PreICH and mPreICH also discriminated ICH in the broader diagnostic cohorts. In the ischemic stroke-versus-ICH cohort, RACE showed an AUC of 0.746 (95% CI, 0.687-0.805), PreICH an AUC of 0.714 (95% CI, 0.661-0.768), and mPreICH an apparent AUC of 0.835 (95% CI, 0.785-0.884). In the overall stroke-code cohort, RACE showed an AUC of 0.774 (95% CI, 0.719-0.828), PreICH an AUC of 0.717 (95% CI, 0.665-0.768), and mPreICH an apparent AUC of 0.812 (95% CI, 0.763-0.861) for identifying ICH (Table 4).

In RACE-stratified analyses, PreICH and mPreICH retained significant discrimination for ICH across both RACE strata. In the LVO-versus-ICH cohort, mPreICH showed an apparent AUC of 0.779 (95% CI, 0.684-0.874) among patients with RACE <=4 and 0.821 (95% CI, 0.749-0.893) among patients with RACE >4 (Table 4). Candidate mPreICH cutoffs are shown in Supplemental Table 3. In the ischemic stroke-versus-ICH cohort, an mPreICH cutoff >=3 yielded a sensitivity of 67.0%, specificity of 88.4%, positive predictive value of 46.8%, negative predictive value of 94.6%, and positive likelihood ratio of 5.78. In the LVO-versus-ICH cohort, the same cutoff yielded a sensitivity of 67.0%, specificity of 77.8%, positive predictive value of 65.6%, negative predictive value of 78.9%, and positive likelihood ratio of 3.02. Higher mPreICH thresholds increased specificity and positive predictive value at the expense of sensitivity (Supplemental Table 3).

## Discussion

In this prospective stroke-code cohort, we evaluated the original PreICH scale in a cohort independent from the derivation study and confirmed its ability to discriminate ICH, including in the clinically relevant LVO-versus-ICH comparison. We also proposed an exploratory modified PreICH score incorporating additional hemorrhage-oriented variables prospectively recorded before neuroimaging. This modified score showed higher apparent discrimination than the original PreICH scale, suggesting that nausea or vomiting, seizure at onset, and graded impairment of consciousness may add diagnostic information beyond overall stroke severity. Importantly, NIHSS and RACE were higher in patients with ICH than in the overall ischemic stroke population, but they no longer differentiated LVO from ICH when the comparison was restricted to patients with confirmed LVO. This finding is central to the present study: severity-based assessment captures the shared clinical severity of LVO and ICH, but it does not adequately resolve the hemorrhage-versus-LVO distinction. This distinction is clinically relevant because LVO and ICH represent the two diagnostic scenarios with the greatest immediate implications for transport decisions, emergency department preparedness, blood pressure management, anticoagulation reversal, and access to thrombectomy-capable or hemorrhage-focused care.^2,6,7,8,14^

Prehospital stroke triage has largely focused on identifying patients with suspected LVO, particularly since the expansion of endovascular thrombectomy and the development of bypass or mothership strategies. Several severity-based prehospital scales, including RACE, G-FAST, CG-FAST, and others, have been developed to enrich for LVO probability, and prospective comparative studies have shown that some of these scales approach clinician-assessed NIHSS for identifying anterior-circulation LVO.^3,4^ However, these tools were primarily designed to identify severe ischemic stroke syndromes rather than to distinguish LVO from ICH. Our findings support this limitation: RACE discriminated ICH from ischemic stroke in the broader confirmed-stroke cohort, likely because ICH patients had more severe neurological presentations, but it did not discriminate LVO from ICH in the primary diagnostic comparison.

The clinical overlap between LVO and ICH is expected. Both conditions may present with abrupt severe focal deficits, gaze deviation, impaired consciousness, and high stroke severity scores. In our cohort, this overlap was evident in the LVO-versus-ICH comparison, where NIHSS and RACE distributions were similar. This reinforces the concept that severity alone is insufficient to determine stroke subtype. Previous clinical literature has consistently shown that features such as vomiting, headache, seizure, coma, impaired consciousness, and elevated blood pressure increase the likelihood of hemorrhagic rather than ischemic stroke, although none of these findings is sufficiently accurate to replace brain imaging.^11,12^

The present study extends the original PreICH work in several ways.^15^ First, PreICH was evaluated prospectively in a broader stroke-code cohort that included ischemic stroke, ICH, and stroke mimics. Second, the analysis addressed not only the broad ischemic stroke-versus-ICH comparison, but also the clinically relevant thrombectomy-era distinction between confirmed LVO and ICH. Third, additional hemorrhage-oriented clinical variables were prospectively recorded before neuroimaging, allowing an exploratory assessment of whether the original score could be refined by incorporating nausea or vomiting, seizure at onset, and graded impairment of consciousness. This approach is consistent with recent systematic reviews of prehospital ICH prediction models, which have highlighted both the potential diagnostic value of simple clinical variables and the limitations of existing models, including retrospective designs, heterogeneous predictor definitions, exclusion of mimics, limited external validation, and high risk of bias.^13^

The main clinical implication of this study is not that a clinical score can replace neuroimaging, but that hemorrhage-oriented clinical information may improve the quality of pre-alert and triage discussions when used alongside LVO screening scales. In contemporary stroke systems, transport decisions often require balancing the potential benefit of direct transfer to a thrombectomy-capable center against the risk of delaying care for patients without LVO. This issue is particularly important in nonurban or regional systems, where bypassing the closest stroke center may substantially increase transport time. The RACECAT trial did not show an overall functional benefit of direct transport to a thrombectomy-capable center in nonurban patients with suspected LVO, and secondary analyses suggested that bypassing the closest stroke center may be harmful in patients ultimately diagnosed with ICH.^8^ Our findings provide a plausible clinical explanation for part of this challenge: a patient with a high LVO severity score may represent either a thrombectomy candidate or a severe hemorrhagic stroke requiring rapid blood pressure management, anticoagulation reversal when appropriate, neurocritical care, and neurosurgical assessment.

Early stroke-subtype differentiation may also have therapeutic implications. INTERACT4 showed that ambulance-delivered intensive blood pressure reduction in suspected acute stroke had different effects according to final stroke subtype, with signals of benefit in patients ultimately diagnosed with ICH and potential harm in those with ischemic stroke.^14^ These findings do not imply that clinical scores should be used to initiate subtype-specific treatment without imaging confirmation. However, they underscore why better early recognition of a hemorrhage-enriched phenotype may be relevant for future prehospital pathways, targeted research, and emergency department preparedness.

Mobile stroke units can resolve diagnostic uncertainty by providing prehospital neuroimaging, but their availability is limited by cost, logistics, and geographic coverage.^9,10^ In most stroke systems, clinical assessment remains the first information available to guide pre-alert and routing. If externally validated, a tool such as mPreICH could potentially serve as a complementary signal to severity-based LVO scales. For example, a high LVO score combined with a high hemorrhage-oriented score might identify patients in whom pre-alert should emphasize both thrombectomy readiness and hemorrhage preparedness. Conversely, such a score should not be used to deny access to thrombectomy-capable centers, delay established acute stroke pathways, or replace urgent neuroimaging.

mPreICH should be interpreted with caution. It was derived after review of the multivariable results and evaluated within the same dataset; therefore, its performance represents apparent discrimination rather than validated diagnostic accuracy. The purpose of the exploratory refinement was not to create a definitive triage rule, but to test whether prospectively recorded variables supported by prior evidence could improve the hemorrhage-oriented signal of the original PreICH scale. The final mPreICH structure is clinically coherent: it preserves systolic blood pressure >160 mm Hg and headache from the original score, adds nausea or vomiting and seizure at onset, and replaces a binary severe-consciousness item with graded weighting of level of consciousness. Diabetes mellitus and anticoagulation were not incorporated because, although they differed in the ischemic stroke-versus-ICH descriptive comparison, they did not distinguish LVO from ICH and were not part of the predefined hemorrhage-oriented clinical construct.

The mPreICH cutoff analyses should be interpreted descriptively. Lower thresholds favored sensitivity and negative predictive value, whereas higher thresholds increased specificity and positive predictive value at the expense of sensitivity. This tradeoff is important in stroke triage, where a high-specificity hemorrhage-oriented signal might be useful to trigger hemorrhage preparedness, but excessive reliance on any clinical score could misclassify patients and delay appropriate treatment. External validation should therefore assess not only discrimination, but also calibration, clinical utility, reproducibility of individual variables, and the consequences of different thresholds in real-world transport models.

This study has several strengths. It was based on consecutive stroke-code activations and included the full diagnostic spectrum of acute stroke-code evaluation, including ischemic stroke, ICH, and stroke mimics. Clinical variables were prospectively recorded by vascular neurologists during the initial evaluation before neuroimaging, reducing the risk that knowledge of imaging findings influenced clinical variable ascertainment.

PreICH was calculated prospectively, allowing direct validation of the original score. RACE was recorded with individual item-level data, enabling detailed analysis of how severity-scale components behave across diagnostic groups. Finally, the study specifically examined the LVO-versus-ICH comparison, which is the most relevant diagnostic challenge for thrombectomy-era triage.

Several limitations should be acknowledged. First, this was a single-center study conducted in a regional stroke system with a high level of vascular neurology involvement at hospital arrival. The findings may not generalize directly to other systems, particularly those in which the initial assessment is performed exclusively by paramedics or emergency medical technicians in the prehospital setting. Second, although variables were recorded before neuroimaging, most assessments occurred at hospital arrival rather than exclusively in the ambulance. Therefore, the results support early stroke-code assessment but do not constitute definitive validation of mPreICH as a prehospital scale. Third, mPreICH was derived and evaluated in the same dataset, and its apparent improvement over PreICH may be overestimated. Fourth, the number of ICH events, although comparable to the original PreICH derivation study, limits the precision of some subgroup and multivariable estimates, as reflected by wide confidence intervals for some predictors. Fifth, variables such as headache, nausea or vomiting, and seizure at onset may be difficult to ascertain in patients with aphasia, decreased consciousness, unwitnessed onset, or limited collateral information. Finally, the study focused on discrimination and candidate operating characteristics; formal calibration, decision-curve analyses, and evaluation of effects on routing decisions, treatment times, and clinical outcomes were not performed.

### Conclusions

In a prospective stroke-code cohort, NIHSS and RACE identified patients with more severe presentations but did not distinguish LVO from ICH. The original PreICH scale showed significant discrimination for ICH, and an exploratory modified version incorporating additional hemorrhage-oriented variables improved apparent discrimination, including in the LVO-versus-ICH comparison. mPreICH should be considered hypothesis-generating and requires external validation before use in prehospital triage or clinical decision-making. Clinical scores may complement early stroke-code assessment and improve hemorrhage-focused preparedness, but neuroimaging remains mandatory for definitive stroke subtype diagnosis.

## Non-standard Abbreviations and Acronyms

ICH: Intracerebral hemorrhage
LVO: Large-vessel occlusion
mPreICH: Modified PreICH scale
NIHSS: National Institutes of Health Stroke Scale
PreICH: Prehospital ICH scale
RACE: Rapid Arterial oCclusion Evaluation

## Author Contributions

FP conceived the study. FP and GM designed the study. AF, GM, YG, AGD, MP, ER, SS, CP, AS, ES, D, and ES contributed to cohort recruitment and clinical data acquisition. RM and ES reviewed neuroimaging. FP and AF performed data processing and data analysis. FP, GM and AF participated in data interpretation and drafted the manuscript. All authors critically revised and approved the final version of the manuscript. FP procured funding.

## Data Availability

Requests for access to the data reported in this article will be considered by the corresponding author.

## Use of Artificial Intelligence

During preparation of this manuscript, the authors used ChatGPT, an artificial intelligence tool developed by OpenAI, solely to assist with English-language editing and improvement of linguistic clarity. The tool was not used for study design, data analysis, data interpretation, generation of scientific content, or formulation of the study conclusions. The authors reviewed and approved all AI-assisted edits and take full responsibility for the final content of the manuscript.

